# Household insecticide use in Amazonian riverine communities: a population-based cross-sectional survey in Belém, Brazil

**DOI:** 10.64898/2026.03.30.26349772

**Authors:** Juliana S. Duarte, Gabriela M. Pereira, Isabelle J. W. Oliveira, Simoneide S. Titze-de-Almeida, Artur F. S. Schuh, Carlos R. M. Rieder, Guilherme T. Valença, Pedro R. P. Brandão, Lane V. Krejcova, Bruno L. Santos-Lobato

**Author notes:** Corresponding Author: Bruno Lopes Santos-Lobato, Laboratório de Neuropatologia Experimental Universidade Federal do Pará Rua dos Mundurucus, 4487, CEP: 66073-000 - Belém, Pará, Brasil.

## Abstract

**Background:** Household insecticides are widely used for domestic pest control, yet exposure patterns in traditionally underserved populations remain poorly characterized. In the Brazilian Amazon, data on use patterns among older adults living in riverine communities are particularly scarce.

**Objective:** To describe the prevalence, frequency, duration, application practices, and types of household insecticides used by older adults living in near-urban riverine insular communities in the Brazilian Amazon.

**Methods:** Cross-sectional, population-based door-to-door survey conducted from August 2022 to July 2025 in four islands (Cotijuba, Mosqueiro, Outeiro, and Combu) in the city of Belém, Brazil. All residents aged 60 years or more registered in the primary care system were invited to participate. Trained interviewers administered an in-person standardized questionnaire to participants on current household insecticide use, frequency, duration, self-application, protective equipment, insecticide types, and product brands.

**Results:** Among 1,101 screened individuals, 1,084 were included (median age at evaluation: 68 years). Overall, 78.4% reported current use of household insecticides. Weekly or more frequent use was reported by 58.9%, and 33.4% reported use for more than 5 years. Self-application was common (57.5%), whereas use of protective equipment was rare (8.2%). Aerosol sprays were the most frequently reported type (39.4%). Commonly recalled aerosols contained pyrethroid mixtures including cypermethrin, imiprothrin, prallethrin, and transfluthrin. A substantial proportion of participants reported using unregulated products and veterinary-only insecticides for household purposes.

**Conclusions:** Household insecticide use is highly prevalent and frequent in Amazonian riverine communities, with minimal use of protective equipment and substantial irregular practices, underscoring the need for targeted risk communication and surveillance.

## 1. Introduction

Pesticide use for occupational and household purposes is increasing worldwide, and 66% of these substances are commercialized for agriculture, while the home and garden sectors account for 24% (World Health Organization and Food and Agriculture Organization of the United Nations, 2020; Grube et al., 2004). Conceived as lethal to target pests, pesticides are also toxic to non-target organisms, including humans, causing acute and chronic adverse effects on human health. Higher prevalences of cancer and neurodegenerative disease are being associated with long-term exposure to pesticides (Galdiano et al., 2021). Human exposure may occur through contaminated soil, air, water, and food.

For this reason, human exposure to pesticides in occupational settings may pose a greater public health risk than in household use. However, household pesticide exposure warrants specific attention for at least three reasons: (I) while the global number of agricultural workers is close to 1 billion, approximately 4.4 billion people live in cities worldwide, and a large proportion of urban residents reported having used pesticides in their lifetime; (II) the informal and unregulated nature of the household pesticides market, together with a limited knowledge on application methods, compared to occupational use, turns this modality of pesticide more hazardous for vulnerable communities; (III) cities are particularly exposed to the effects of ongoing climate changes, increasing the challenge of urban pest control (Santos-Lobato, 2025).

Therefore, the impacts of household pesticide exposure on human health may be comparable in relevance to those of occupational exposure.

Insecticides are the most commonly used household pesticides, accounting for around 50% of household pesticides commercialized (Grube et al., 2004). Paradoxically, few studies examine patterns of household insecticide use across diverse populations. Most evidence comes from cross-sectional surveys conducted in high-income countries and selected urban settings in middle-income regions, with limited information on household insecticide use in remote or traditionally underserved populations, where housing conditions, pest burden, access to regulated products, and risk communication may differ markedly from those in large urban centers. To address this gap, we conducted a population-based, door-to-door survey of older adults residing in riverine communities in the Brazilian Amazon. To date, no population-based surveys have assessed household insecticide use in riverine communities in the Brazilian Amazon, particularly among the potentially more vulnerable group to the cumulative effects of such exposure, the older adults.

## 2. Methods

### 2.1. Study design and participants

This study is part of the Prodromal and Overt Parkinson’s Disease Epidemiological Study in Brazil (PROBE-PD), a research project investigating the prevalence of parkinsonism and its risk factors across four Brazilian regions (NCT05677529). We conducted a cross-sectional, observational, population-based study to examine the pattern of household insecticide use in near-urban, riverine, traditional communities in Belém, the main city of the Brazilian Amazon.

We contacted all individuals aged 60 or over who were registered with the primary care of the public health system in four insular communities: Cotijuba, Mosqueiro, Outeiro, and Combu Islands (Fig. 1) through door-to-door assessments from August 2022 to July 2025. The approach aimed to achieve a census-based assessment for the target group population. Mosqueiro and Outeiro are classified as more urbanized places based on population density and infrastructure. The study was approved by the Ethics Committee of the Health Sciences Institute of the Universidade Federal do Pará (Number 7.164.572), and all participants provided written informed consent.

**Fig 1.**
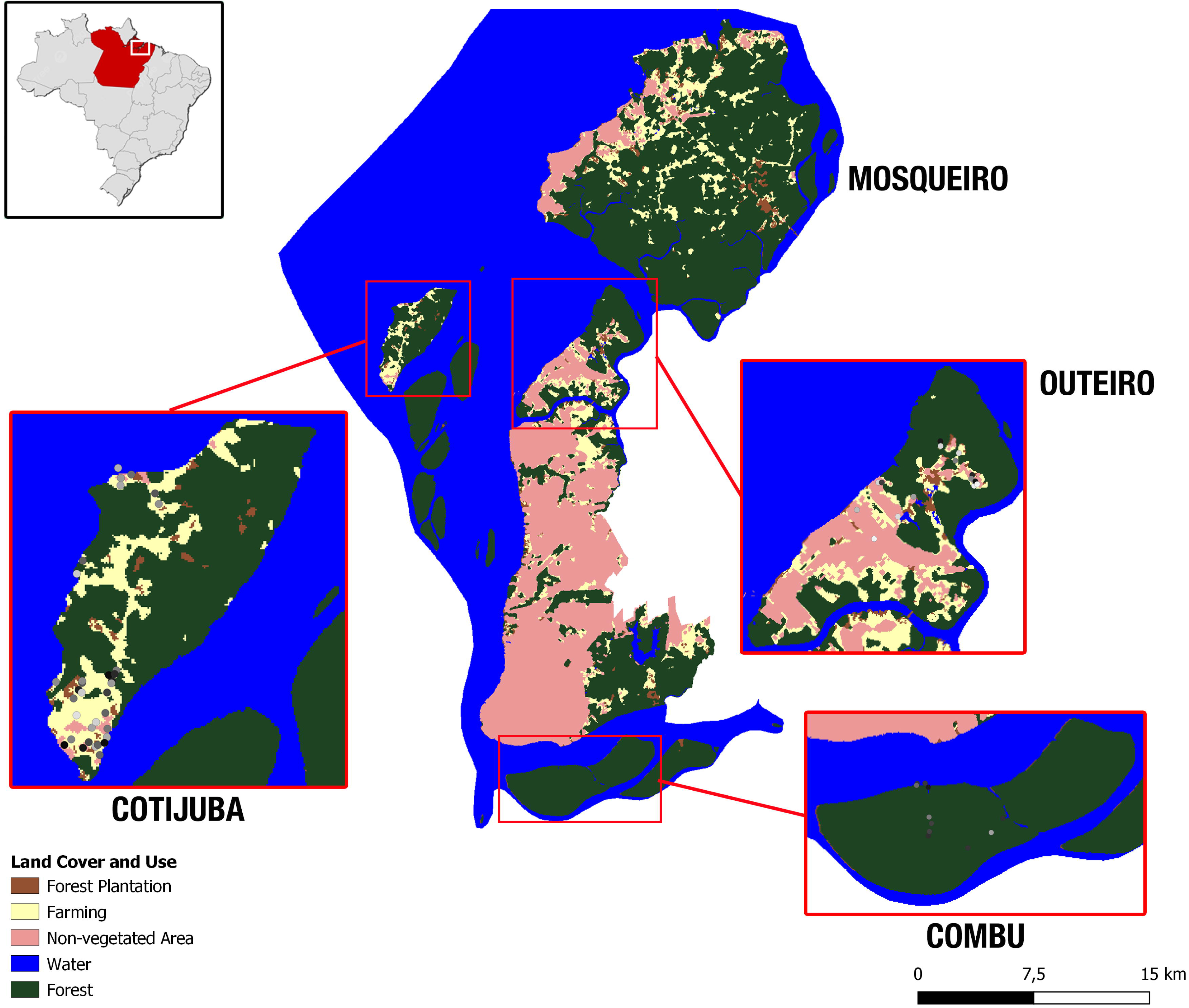

### 2.2. Household insecticide use survey

All participants assessed during the door-to-door approach were interviewed in person about their household insecticide use. We defined household insecticide as any product used in or around the home (e.g., in the garden or on the lawn) to control ants, mosquitoes, cockroaches, termites, and other insects, excluding treatments for head lice or pet parasites. We used a standard questionnaire (in Portuguese) to collect information on their methods for killing insect pests. The full questionnaire is available as supplementary online material in the original Portuguese version and its English translation (Supplementary Material S1).

The initial question was whether the participant currently used any chemical product to kill insects at the time of the interview. If the answer was No, the questionnaire was concluded. If the answer was Yes, further questions were made focusing on the frequency of use (more than once a week, once a week, once per month, once every three months, every six months, once per year); duration of use (less than 5 years, between 5 and 10 years, between 10 and 15 years, between 15 and 20 years, between 20 and 30 years, and over 30 years); whether the participant personally applied insecticides and if used any personal protective equipment.

We also asked about the types of insecticides used to control pests (aerosol spray cans, electric plug-in emitters, mosquito coils, baits, other types) and the product brands. To ensure that participants correctly recalled the brand varieties of consumer products, the questionnaire presented color images of insecticide types, including four main local insecticide aerosol spray can brands, and responses were recorded using a multiple-choice format. Active ingredients and concentration ranges were obtained from the manufacturers’ safety data sheets for representative aerosol formulations of the brands shown during the interview, to describe the most common pyrethroid mixtures and concentration ranges. Active ingredient toxicity was classified according to the WHO Recommended Classification of Pesticides by Hazard (WHO Hazard) and the Globally Harmonized System of Classification and Labelling of Chemicals (GHS) for oral exposure in acute human health (Supplementary Table S1) (World Health Organization, 2020).

### 2.3. Statistical analysis

Data collected were entered into REDCap (Research Electronic Data Capture) (Harris et al. 2009) and exported to Microsoft Excel before data analysis. To compare frequencies across categories, we used the chi-square test, and to compare medians, we used the Kruskal-Wallis test. Missing data were not imputed. Statistical significance was set at p < 0.05 (two-tailed). All analyses were performed using SPSS for Windows version 23.0 (SPSS Inc., Chicago, USA).

## 3. Results

We screened 1,101 individuals aged 60 or older and, after excluding 17 participants with incomplete questionnaire responses, selected 1,084 participants for this study. Demographic data are summarized in Table 1.

**Table 1.**
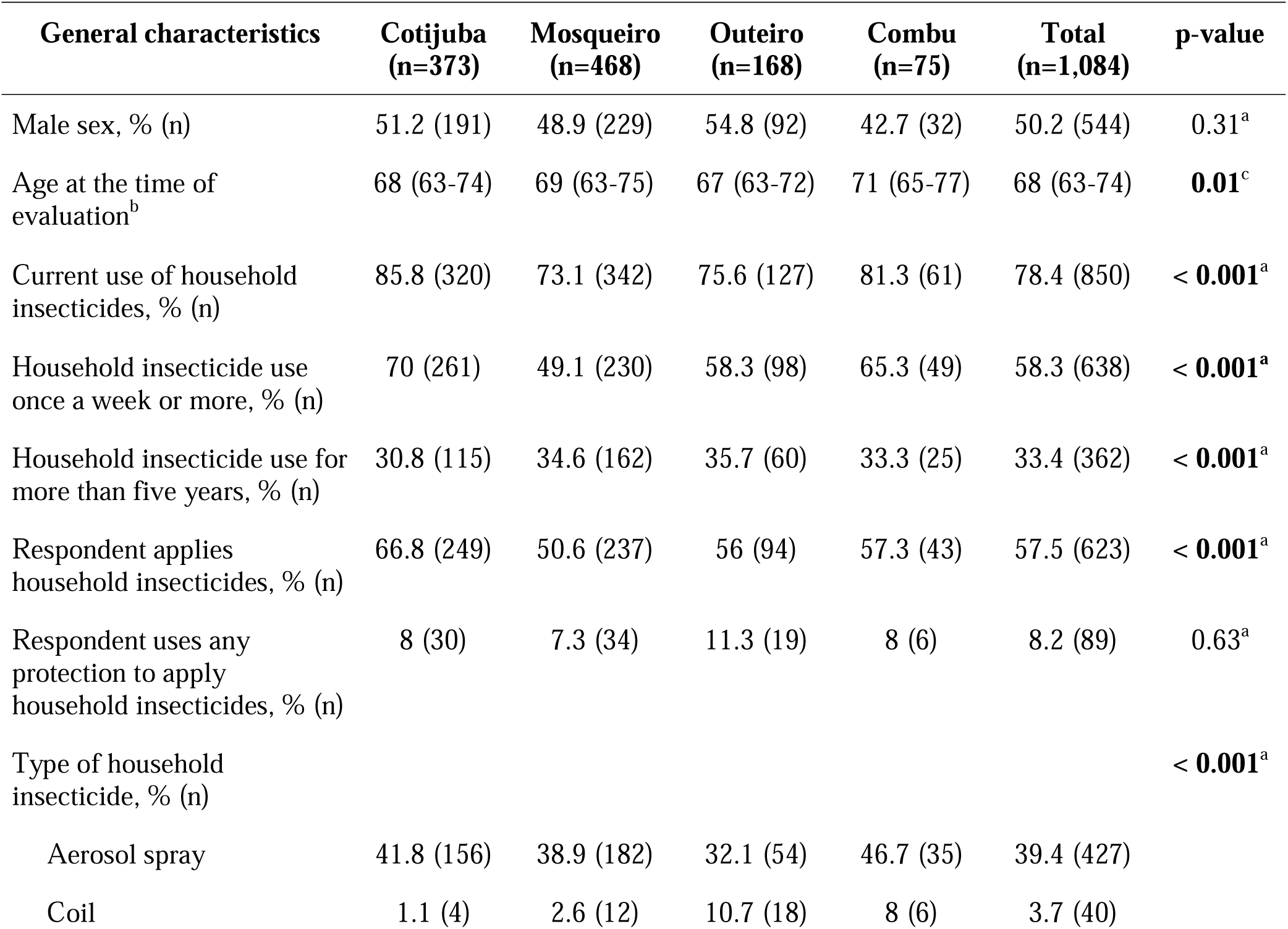
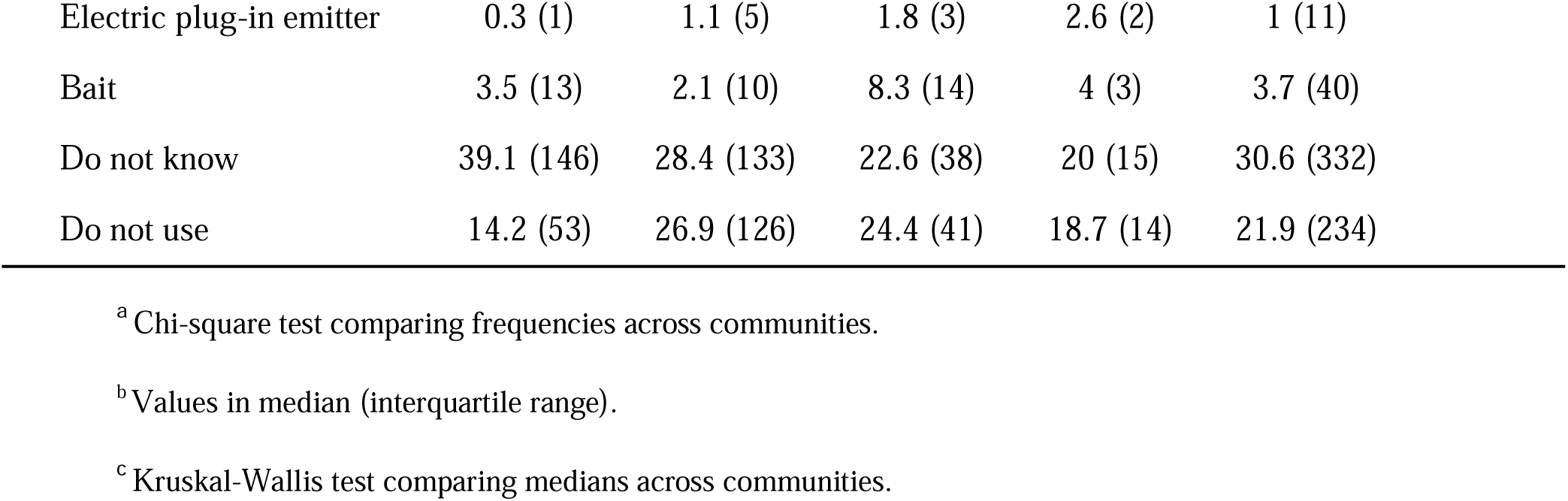
Demographic data of participants according to the riverine community.

| **General characteristics** | **Cotijuba**<br>**(n=373)** | **Mosqueiro**<br>**(n=468)** | **Outeiro**<br>**(n=168)** | **Combu**<br>**(n=75)** | **Total**<br>**(n=1,084)** | **p-value** |
| --- | --- | --- | --- | --- | --- | --- |
| Male sex, % (n) | 51.2 (191) | 48.9 (229) | 54.8 (92) | 42.7 (32) | 50.2 (544) | 0.31<sup>a</sup> |
| Age at the time of<br>evaluation<sup>b</sup> | 68 (63-74) | 69 (63-75) | 67 (63-72) | 71 (65-77) | 68 (63-74) | **0.01<sup>c</sup>** |
| Current use of household<br>insecticides, % (n) | 85.8 (320) | 73.1 (342) | 75.6 (127) | 81.3 (61) | 78.4 (850) | **< 0.001<sup>a</sup>** |
| Household insecticide use<br>once a week or more, % (n) | 70 (261) | 49.1 (230) | 58.3 (98) | 65.3 (49) | 58.3 (638) | **< 0.001<sup>a</sup>** |
| Household insecticide use for<br>more than five years, % (n) | 30.8 (115) | 34.6 (162) | 35.7 (60) | 33.3 (25) | 33.4 (362) | **< 0.001<sup>a</sup>** |
| Respondent applies<br>household insecticides, % (n) | 66.8 (249) | 50.6 (237) | 56 (94) | 57.3 (43) | 57.5 (623) | **< 0.001<sup>a</sup>** |
| Respondent uses any<br>protection to apply<br>household insecticides, % (n) | 8 (30) | 7.3 (34) | 11.3 (19) | 8 (6) | 8.2 (89) | 0.63<sup>a</sup> |
| Type of household<br>insecticide, % (n) |  |  |  |  |  | **< 0.001<sup>a</sup>** |
| Aerosol spray | 41.8 (156) | 38.9 (182) | 32.1 (54) | 46.7 (35) | 39.4 (427) |  |
| Coil | 1.1 (4) | 2.6 (12) | 10.7 (18) | 8 (6) | 3.7 (40) |  |

|  |  |  |  |  |  |
| --- | --- | --- | --- | --- | --- |
| Electric plug-in emitter | 0.3 (1) | 1.1 (5) | 1.8 (3) | 2.6 (2) | 1 (11) |
| Bait | 3.5 (13) | 2.1 (10) | 8.3 (14) | 4 (3) | 3.7 (40) |
| Do not know | 39.1 (146) | 28.4 (133) | 22.6 (38) | 20 (15) | 30.6 (332) |
| Do not use | 14.2 (53) | 26.9 (126) | 24.4 (41) | 18.7 (14) | 21.9 (234) |

<sup>a</sup> Chi-square test comparing frequencies across communities.
<sup>b</sup> Values in median (interquartile range).
<sup>c</sup> Kruskal-Wallis test comparing medians across communities.

We found that 78.4% of participants reported using household insecticides. The proportion was lower in more urbanized communities (Mosqueiro, 73.1%; Outeiro, 75.6%; p < 0.001). Based on frequency and duration of use, 58.9% of participants reported using household insecticides once a week or more, and 33.4% reported using the products for more than 5 years; lower frequency and longer duration of use were more common in communities with higher urbanization (p < 0.001). 57.5% of participants reported applying household insecticides themselves, but only 8.2% used any personal protective equipment. Aerosol spray was the most used type of household insecticide (39.4%) (Fig. 2).

**Fig 2.**
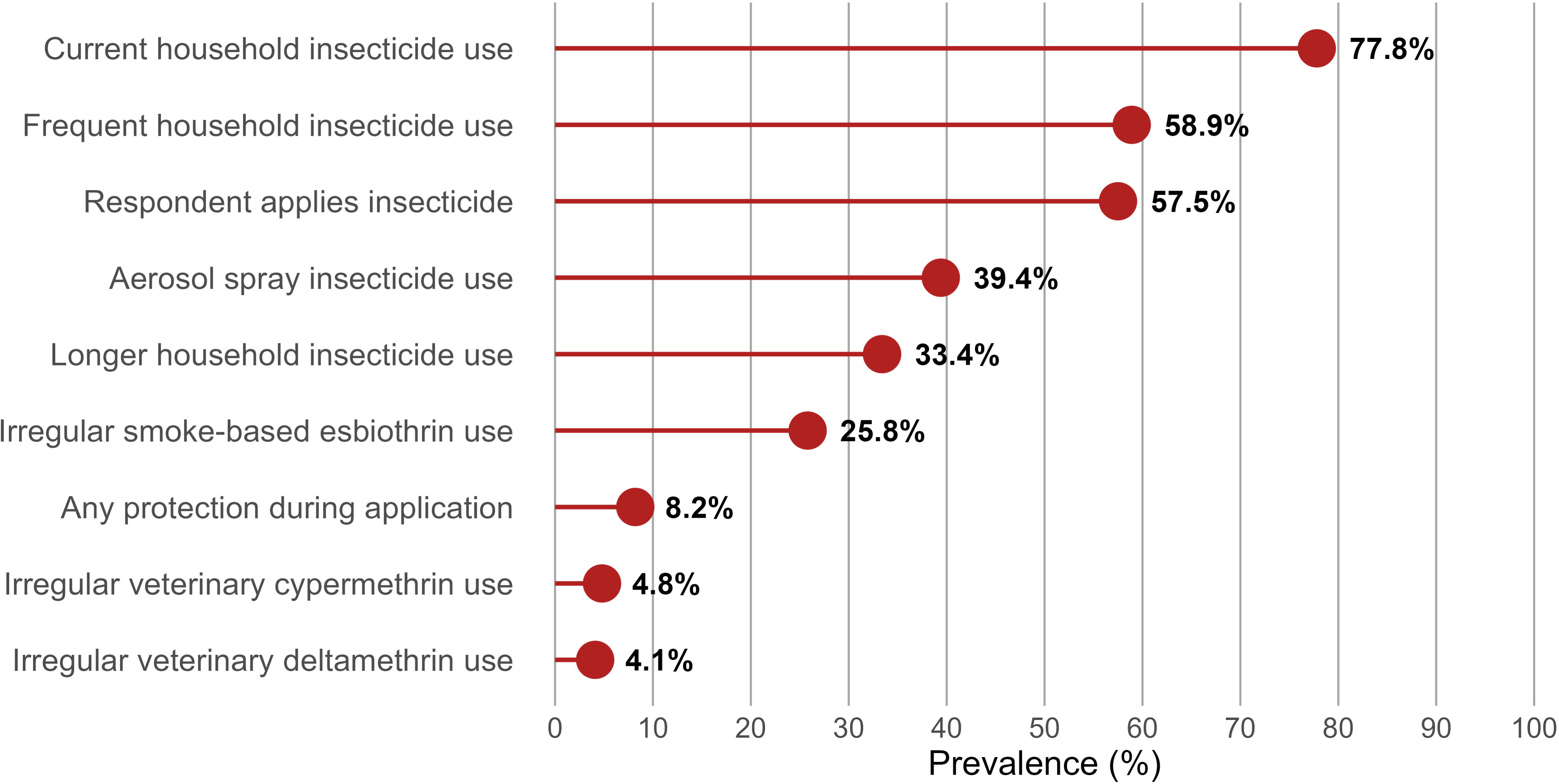

As the most commonly used type of household insecticide, aerosol sprays were analyzed from four reported brands. All products contained a combination of pyrethroids (brand 1, from manufacturer 1: cypermethrin 0.05-0.15%, prallethrin 0.02-0.05%, imiprothrin 0.04-0.08%; brands 2, 3, and 4, from manufacturer 2: transfluthrin 0.01-0.5%, cypermethrin 0.01-0.5%, imiprothrin 0.01-0.5%) (Supplementary Table S1). Among participants who reported using aerosol sprays, brand 1 was the most frequently recalled household insecticide (81.7%).

Further, respondents spontaneously cited alternative household insecticides. For example, 25.8% of participants reported using a smoke-based insecticide supposedly formulated with esbiothrin (Supplementary Table S1). Most participants described the product as a “natural incense” with no potential hazard. The product has not received approval from the Brazilian Health Regulatory Agency but is freely commercialized in informal markets, including online. Participants also cited the use of two water-emulsifiable concentrate insecticides for veterinary use only: one containing cypermethrin 150 g/L and the other containing deltamethrin 50 g/L (Supplementary Table S1). Participants reported using these products indoors and around the home, either by spraying or direct application of the liquid solution. Despite the instructions on the product label, most people described diluting, applying, and discharging these insecticides without any particular care. Use of these products was reported at 4.8% (cypermethrin 150 g/L) and 4.1% (deltamethrin 50 g/L).

## 4. Discussion

The present study showed that household insecticide use is highly prevalent and frequent, with most users reporting at least weekly application without protective equipment, in riverine communities near a major city in the Brazilian Amazon. The informal commercialization of insecticides was evidenced by the use of an illegal product by one quarter of the sample and the irregular use of veterinary insecticides. These practices suggest potential for chronic exposure and unsafe handling.

Our findings are similar to those reported in previous studies. Most household pesticide surveys in developed regions (Savage et al., 1981; Grey et al., 2006), in Brazilian urban zones (Diel et al., 2003; Oliveira et al., 2015; Galdiano et al., 2021), and in rural areas of other developing countries (Chitra et al., 2013) reported a high prevalence of use exceeding 80%, independent of geographic or climatic differences (Table 2). Among different household pesticide classes, insecticides were the most commonly reported in all studies (Savage et al., 1981; Bass et al., 2001; Diel et al., 2003; Grey et al., 2006; Oliveira et al., 2015; Galdiano et al., 2021; Porusia et al., 2025).

**Table 2.** Characteristics of previous household insecticide use surveys.

| Author and Year | Country | Local setting | Sample size | Male sex (%) | Use (%)<sup>a</sup> | Weekly use or more (%) | Use of PPE (%) | Pyrethroids as main insecticide |
| --- | --- | --- | --- | --- | --- | --- | --- | --- |
| Savage et al., 1981 | USA | Mixed | 8,254 | NA | 90.7 | NA | NA | No |
| Whitmore et al., 1993 | USA | Mixed | 84,573 | NA | NA | NA | 30 | Yes |
| Bass et al., 2001 | USA | Urban | 107 | 15 | 91.2 | 31.7 | NA | Yes |
| Diel et al., 2003 | Brazil | Urban | 2,039 | NA | 89 | NA | NA | Yes |
| Gray et al., 2006 | United Kingdom | Mixed | 713 | NA | 93 | NA** | NA | Yes |
| Chitra et al., 2013 | India | Rural | 143 | 32.2 | 96.5 | 92.2 | NA | NA |
| Oliveira et al., 2015 | Brazil | Urban | 700 | 25 | 85 | NA | 26 | NA |
| Galdiano et al., 2021 | Brazil | Urban | 927 | 32.4 | 87.5 | NA | NA | Yes |
| Kim et al.,<br>2021 | South<br>Korea | Mixed | 1,239 | 30.4 | NA | 12.7 | NA | NA |
| Present study | Brazil | Mixed | 1,084 | 50.2 | 78.4 | 58.9 | 8.2 | Yes |

Abbreviations: NA, not available; PPE, personal protective equipment.
\* Prevalence of household insecticide use (current or during the last months).
\*\* Data not available; however, the median annual frequency of use for insecticides at home was 3 (interquartile range 1-10).

As demonstrated by our results, aerosol sprays are the predominant insecticide application method (Whitmore et al., 1993; Diel et al., 2003; Grey et al., 2006; Oliveira et al., 2015; Galdiano et al., 2021), except among people living in rural South India, who primarily use mosquito coils (Chitra et al., 2013). Furthermore, a direct correlation between the use of aerosol sprays and household income was observed, and more economically vulnerable households reported greater use of pulverized insecticides or smoke-based mosquito coils (Diel et al., 2003), suggesting that economic and cultural factors shape the use of household insecticides. Pyrethroids were the most commonly used insecticide class in the most recent studies (Bass et al., 2001; Diel et al., 2003; Grey et al., 2006; Galdiano et al., 2021), as reported in the present study. However, older surveys from the 1980s in the USA reported that dichlorvos, a highly toxic organophosphate, was the most widely used household insecticide in the country (Savage et al., 1981). One decade later, pyrethroids were already the most prevalent household insecticide in the USA (Whitmore et al., 1993). The use of irregular household insecticides was also detected by previous studies, through banned products such as the “Miraculous Insecticide Chalk” or “poison chalk”, used in the USA and India (Bass et al., 2001; Chitra et al., 2013).

Regarding the frequency of use, a previous study from the USA showed that 30% of participants described using insecticides once a week or more (Bass et al., 2001), while in a report from the United Kingdom, household insecticides were used in a median of three times per year (Grey et al., 2006). Only 12% of elders aged 60 or more from South Korea reported applying insecticides once a week or more (Kim et al., 2021). Compared to these results from developed regions, the prevalence of participants who used household insecticides once a week or more in the present study (58.9%) is substantially higher. Results from environmental questionnaires may be correlated with insecticide exposure levels, because insecticide use frequency is associated with urinary concentrations of 3-PBA, the main pyrethroid metabolite used to assess pyrethroid exposure (Kim et al., 2021). More concerning is the lack of safety precautions during insecticide application, with fewer than 10% of participants reporting the use of any protective equipment. Another Brazilian survey also reported a low frequency (26%) of protective equipment use when applying insecticides (Oliveira et al., 2015). Still, the high prevalence of knowledge about the need to use protective equipment when applying and mixing pesticides (80.4% and 86.2%, respectively) in South Asia with higher education (Porusia et al., 2025) suggests that these practices may be associated with cultural and educational backgrounds.

Although not evaluated in our study, some patterns of household insecticide use have been previously described. Most people applied the products inside their homes, mainly in bedrooms, living rooms, kitchens, and bathrooms (Whitmore et al., 1993; Bass et al., 2001; Chitra et al., 2013; Galdiano et al., 2021; Porusia et al., 2025). Mosquitoes, ants, and cockroaches were the most frequently reported pests in their homes (Whitmore et al., 1993; Chitra et al., 2013; Galdiano et al., 2021; Porusia et al., 2025). Many respondents reported that they acquired insecticides even without an active pest invasion in their homes, indicating a preventive behavior of storing for use when needed (Bass et al., 2001; Diel et al., 2003; Chitra et al., 2013; Galdiano et al., 2021; Porusia et al., 2025). Unsafe practices may substantially increase exposure to pesticides. In South India, among mosquito coil users, 61% reported keeping doors and windows closed while the coil was burning, and nearly half did not recognize household pesticides as harmful to their own health or to children’s health (Chitra et al., 2013). This illustrates how perceived “routine” pest-control products may generate avoidable exposure in poorly ventilated indoor environments.

Together, evidence indicates a rising use of household insecticides worldwide for mosquito and other pest control, particularly pyrethroids, in urban and rural settings, including riverine communities in the Brazilian Amazon. Pyrethroids have been the most widely used insecticides globally since the 1980s, owing to their perceived low toxicity and the abandonment of more toxic pesticide classes, such as organophosphates. In 2023, the pyrethroid market was valued at US$ 4.2 billion, with a projected future growth to US$ 6.8 billion in 2032 due to increasing agricultural activities, rising awareness of insect-borne diseases, rapid urbanization and industrial development leading to a higher demand for pest control, and ongoing advancements in formulation technologies and product innovations (Global Market Insights, 2024). In the Brazilian market, all insecticide formulations increased from 2022 to 2024, with aerosol sprays leading, accounting for 2.7 billion Brazilian reais in 2024 (APIPLA, 2025).

The broader context in the Brazilian Amazon is characterized by historical and ongoing reliance on insecticides for mosquito borne disease control. Yellow fever and malaria caused multiple outbreaks in the region during the colonial and imperial periods. Following the acceptance of the mosquito borne disease theory in the early 20th century, public health strategies focused on eliminating breeding sites near urban areas. During World War II, when the city of Belém hosted Allied troops, health authorities and the U.S. military implemented intensive pest control programs, including the first large scale use of DDT in the region (Campos, 2006). DDT was later adopted as the main insecticide for mosquito control in Brazil and contributed to the temporary eradication of Aedes aegypti in 1955 (Alsharif et al., 2025). Subsequent resurgences of Aedes aegypti and the emergence of insecticide resistance led to the introduction of organophosphates (e.g., temephos, malathion) and then pyrethroids (Alsharif et al., 2025). Recent epidemics of dengue, chikungunya, Zika, and Oropouche fever have further reinforced reliance on insecticides.

This historical trajectory may have normalized routine household insecticide storage and use in tropical regions (Chitra et al., 2013), potentially contributing to the high weekly use observed in our study. Current Brazilian vector control policies combine multiple low dose insecticide classes (pyrethroids, carbamates, neonicotinoids) with Wolbachia infected mosquitoes to mitigate resistance (Alsharif et al., 2025). Nevertheless, resistance dynamics often drive increases in dose and application frequency over time, suggesting that the global circulation of pyrethroids is likely to grow.

Consistent with this, the global circulation of pyrethroids has been associated with increased detection of pyrethroid residues in various media, including soils, surface water, sediments, crops, aquatic organisms, and indoor environments, as well as in human biological samples (urine and human milk) (Tang et al., 2018). In particular environmental conditions, the degradation of pyrethroids applied in agricultural and household settings occurs partially. These substances are transported to streams and rivers via soil, may reach groundwater and the marine environment, and bioaccumulate in fruits, vegetables, and fish (Tang et al., 2018). Given this, previous studies have reported that inadequate disposal of household insecticides is common (Bass et al., 2001; Chitra et al., 2013). The scenario is compatible with global pyrethroid pollution.

Pyrethroid household insecticides are often perceived as relatively “low toxicity” consumer products when compared with older classes such as organophosphates. This misperception may be explained by the fact that most hazard classifications for pesticides are based solely on acute health hazard (i.e., over a relatively short period), such as the WHO Hazard and the GHS categories. According to this classification, most pyrethroid active ingredients are classified as Class II (moderately hazardous) (World Health Organization, 2020). To harmonize risk classifications, the United Nations proposed the GHS, which classifies most pyrethroids as Category 4 (moderate toxicity) for oral exposure in acute human health (World Health Organization, 2020). Neither classification accounts for chronic effects on human health, indicating that these classifications fail to capture the negative impacts of long-term, low-dose pyrethroid exposure. This is a relevant limitation, because multiple chronic diseases and toxic effects have been associated with pyrethroid exposure in observational studies, including different cancer types, male infertility, and cardiovascular disease (Tang et al., 2018; Galdiano et al., 2021). For example, Parkinson’s disease is one of the main diseases associated with pesticide exposure (Silva et al., 2026), and household insecticide use may increase the risk of Parkinson’s disease by 2-fold (Moura et al., 2023; Santos-Lobato et al., 2025). Furthermore, older people may be more vulnerable to pesticides due to cumulative exposure over their lifetimes (Tang et al., 2018).

As strong points, to the best of our knowledge, this is the first survey of household insecticide use in the Amazon. Additionally, the present study employed a door-to-door approach and in-person interviews to administer the standard questionnaire, achieving high coverage of target communities. The evaluated traditional populations may represent patterns of household insecticide use in most riverine communities in the Brazilian Amazon and other tropical forest regions. As limitations, our small sample size precluded regression analyses to estimate the association between insecticide use and PD, and the study focused on a strict age range (60 years or more), which reduces the generalizability of the results, even when selecting a population vulnerable to the exposure. Furthermore, the cross-sectional design of the study precludes causal inference, and exposure was assessed using a self-report instrument, which may be subject to misclassification and recall bias. Additionally, pesticide exposure was quantified using questionnaires, which are susceptible to heterogeneity and recall bias, and our environmental questionnaire did not examine socioeconomic factors or other behaviors associated with insecticide use.

## 5. Conclusions

In conclusion, although household insecticide use patterns in the Brazilian Amazon are similar to those described globally, the frequency of use in these communities was significantly higher than in other regions, including other regions of Brazil. These findings underscore the importance of monitoring pyrethroid pollution, even in purportedly pristine ecological sanctuaries and environmentally sensitive regions, such as the Amazonian rainforest. The long-term effects of this exposure on human health remain unknown, but evidence suggests numerous adverse impacts. Thus, actions such as promoting targeted risk communication, embedding safer-use interventions in primary care and community health work, and strengthening surveillance and regulatory measures to address irregular products are needed. Future studies incorporating biomonitoring and indoor/environmental measurements in Amazonian settings are warranted to quantify better chronic exposure and its potential health implications in older populations.

## CRediT authorship contribution statement

**Juliana S. Duarte:** Writing – review & editing, Supervision, Methodology, Formal analysis, Data curation, Investigation, Conceptualization. **Gabriela M. Pereira:** Writing – review & editing, Methodology, Investigation, Project administration, Conceptualization. **Isabelle J. W. Oliveira:** Writing – review & editing, Methodology, Investigation. **Simoneide S. Titze-de-Almeida:** Writing – review & editing, Methodology, Investigation. **Artur F. S. Schuh:** Writing – review & editing, Funding acquisition, Methodology, Investigation, Conceptualization. **Carlos R. M. Rieder:** Writing – review & editing, Methodology, Investigation, Conceptualization. **Guilherme T. Valença:** Writing – review & editing, Methodology, Investigation, Conceptualization. **Pedro R. P. Brandão:** Writing – review & editing, Methodology, Investigation, Conceptualization. **Lane V. Krejcova:** Writing – review & editing, Supervision, Methodology, Data curation, Investigation, Conceptualization. **Bruno L. Santos-Lobato:** Writing – original draft, Writing – review & editing, Supervision, Methodology, Formal analysis, Data curation, Investigation, Conceptualization.

## Acknowledgements

We would like to thank all participants in PROBE-PD, as well as the staff at the Brazilian sites, for their efforts.

## Data availability

Data will be made available on request.

## References

Alsharif, B., Melo-Santos, M.A.V., Barbosa, R.M.R., Junqueira Ayres, C.F., 2025. A Brief History of the Use of Insecticides in Brazil to Control Vector-Borne Diseases, and Implications for Insecticide Resistance. Trop. Med. Infect. Dis. 10. 10.3390/tropicalmed10120336

Associação Brasileira das Indústrias de Produtos de Higiene, Limpeza e Saneantes (ABIPLA). Anuário ABIPLA 2025. 20th ed. ABIPLA, São Paulo, Brazil.

Bass, J.K., Ortega, L., Rosales, C., Petersen, N.J., Philen, R.M., 2001. What’s being used at home: a household pesticide survey. Rev. Panam. Salud Publica 9, 138–144.

Campos, A.L.V., 2006. O controle de malária nas bases militares norteamericanas no Brasil, in: Políticas Internacionais de Saúde na Era Vargas: O Serviço Especial de Saúde Pública, 1942-1960. FIOCRUZ, pp. 91–112.

Chitra, G.A., Kaur, P., Bhatnagar, T., Manickam, P., Murhekar, M.V., 2013. High prevalence of household pesticide use and unsafe use in rural South India. Int. J. Occup. Med. Environ. Health 26, 275–282.

Diel, C., Facchini, L.A., Dall’Agnol, M.M., 2003. [Household insecticides: pattern of use according to per capita income]. Rev. Saude Publica 37, 83–90.

Galdiano, L.L.S., Baltar, V.T., Polidoro, S., Gallo, V., 2021. Household pesticide exposure: an online survey and shelf research in the Metropolitan Region of Rio de Janeiro, Brazil. Cad. Saude Publica 37, e00099420.

Global Market Insights. Pyrethroids market size, share & forecast report, 2024–2032. Report ID: GMI8841. https://www.gminsights.com/industry-analysis/pyrethroids-market (accessed February 2026).

Grey, C.N.B., Nieuwenhuijsen, M.J., Golding, J., ALSPAC Team, 2006. Use and storage of domestic pesticides in the UK. Sci. Total Environ. 368, 465–470.

Grube, A.H., Donaldson, D., Kiely, T., 2004. Pesticides Industry Sales and Usage: 2000 and 2001 Market Estimates, Environmental Protection Agency (USA), Biological and Economic Analysis Division.

Kim, J.H., Kim, S., Hong, Y.C., 2021. Household insecticide use and urinary 3-phenoxybenzoic acid levels in an elder population: a repeated measures data. J. Expo. Sci. Environ. Epidemiol. 31, 1017–1031.

Moura, D.D., Borges, V., Ferraz, H.B., Schuh, A.F.S., de Mello Rieder, C.R., Mata, I.F., Brito, M.M.C.M., Tumas, V., Santos-Lobato, B.L., 2023. History of high household pesticide use and Parkinson’s disease in Brazil. Parkinsonism Relat. Disord. 113, 105493.

Porusia, M., Chotklang, D., Mohd Yatim, S.R., Purnamasari, S., Ramadhani, D.S., Cahyani, P., Nabila, K.N., Aulia, A.Z., Mokhtar, A.S., Low, V.L., Sahimin, N., 2025. Household pesticide use in Southeast Asia: a comparative survey of knowledge, attitudes, and practices in Indonesia, Thailand and Malaysia. J. Integr. Pest Manag. 16, pmaf039.

Oliveira, L.B., Nunes, R.M.P., Santana, C.M., Costa, A.R., Nunes, N.M.F., Calou, I.B.F., Peron, A.P., Marques, M.M.M., Ferreira, P.M.P., 2015. Profile of the population use of household insecticides against mosquitoes. Semina: Ciências Biológicas e da Saúde 36, 79–92.

Santos-Lobato, B.L., 2025. New Insights into the Association of Pesticide Exposure and Parkinson’s Disease. Mov. Disord. 40, 579–580.

Santos-Lobato, B.L., Schuh, A.F.S., Mata, I.F., 2025. Household herbicide use is associated with the risk of Parkinson’s disease and a younger age at onset in PPMI and Fox Insight cohorts. J Neural Transm (Vienna). 10.1007/s00702-025-03041-8

Savage, E.P., Keefe, T.J., Wheeler, H.W., Mounce, L., Helwic, L., Applehans, F., Goes, E., Goes, T., Mihlan, G., Rench, J., Taylor, D.K., 1981. Household pesticide usage in the United States. Arch. Environ. Health 36, 304–309.

Silva, P.H.P., Abreu, J.S., Gomes, R.S.G., Oliveira, H.K.A., Schuh, A.F.S., Mata, I.F., Santos-Lobato, B.L., 2026. Revisiting the Association of Pesticide Exposure and Parkinson’s Disease: Systematic Review and Meta-Analysis. Environ. Toxicol. 10.1002/tox.70041

Tang, W., Wang, D., Wang, J., Wu, Z., Li, L., Huang, M., Xu, S., Yan, D., 2018. Pyrethroid pesticide residues in the global environment: An overview. Chemosphere 191, 990–1007.

Whitmore, R.W., Kelly, J.E., Reading, P.L., Brandt, E., Harris, T., 1993. National home and garden pesticide use survey, in: Racke K.D., Leslie A.R. (Eds), Pesticides in urban environments. American Chemical Society, Washington DC, pp. 18–36.

World Health Organization, 2020. WHO recommended classification of pesticides by hazard and guidelines to classification, 2019 edition. World Health Organization, Geneve.

World Health Organization and Food and Agriculture Organization of the United Nations, 2020. International Code of Conduct on Pesticide Management: guidance on management of the household pesticides. World Health Organization and Food and Agriculture Organization of the United Nations, Geneve.

