## Supplementary Material S1 for "Household insecticide use in Amazonian riverine communities: a population-based cross-sectional survey in Belém, Brazil"

### Supplemental Material

S1-S2. Household insecticide use questionnaire (simplified versions for publication).

#### S1. Questionário (Português)

**Título:** Hábitos domiciliares do uso de inseticidas (uso domiciliar)

**Instruções:** As perguntas abaixo avaliam hábitos de uso de inseticidas no domicílio. Responda de acordo com sua rotina atual.

##### A. Dados gerais

1. **Idade (anos): ________**

2. **Sexo:**

   ( ) Masculino
   ( ) Feminino

3. **Onde mora atualmente:**

   ( ) Belém
   ( ) Ananindeua
   ( ) Marituba
   ( ) Outra cidade do Pará
   ( ) Fora do estado do Pará

##### B. Uso domiciliar de inseticidas

4. **Você usa inseticidas em casa?**

   ( ) Sim
   ( ) Não

*Se NÃO, encerre o questionário.*

5. **Há quanto tempo você faz uso de inseticidas?**

   ( ) Menos que 5 anos
   ( ) Entre 5 e 10 anos
   ( ) Entre 10 e 15 anos
   ( ) Entre 15 e 20 anos
   ( ) Entre 20 e 25 anos
   ( ) Entre 25 e 30 anos
   ( ) Mais que 30 anos

6. **Qual a frequência de uso de inseticidas?**

   ( ) Mais de uma vez por semana
   ( ) Uma vez por semana
   ( ) Uma vez por mês
   ( ) Uma vez a cada 3 meses
   ( ) Uma vez a cada 6 meses
   ( ) Uma vez ao ano

7. **Você é quem aplica o inseticida na sua residência?**

( ) Sim
( ) Não

8. **Ao utilizar o inseticida, você costuma usar alguma proteção (máscara, luva ou outra)?**

( ) Sim
( ) Não
( ) Às vezes

9. **Qual tipo de inseticida você costuma usar com mais frequência?**

( ) Aerossol (spray)
( ) Espiral/bobina (mosquito coil)
( ) Isca
( ) Elétrico (líquido)
( ) Elétrico (pastilha)
( ) Outro: ________

*Nota: no questionário aplicado em campo, foram apresentadas imagens coloridas ilustrativas dos tipos de inseticidas para auxiliar a recordação do participante.*

10. **(Somente se marcou “Aerossol/spray” na pergunta 9)** Entre as opções abaixo, qual marca de aerossol você mais utiliza em sua residência?

( ) Brand 1
( ) Brand 2
( ) Brand 3
( ) Brand 4
( ) Outra: ________

*Nota: no questionário original, foram apresentadas imagens coloridas das embalagens das quatro marcas de aerossol para facilitar a identificação.*

#### S2. Questionnaire (English)

**Title:** Household insecticide use (household setting)

**Instructions:** The questions below assess household insecticide use. Please answer based on your current routine.

##### A. General information

1. **Age (years): ________**

2. **Sex:**

( ) Male
( ) Female

3. **Current place of residence:**

( ) Belém
( ) Ananindeua
( ) Marituba
( ) Another city in the state of Pará
( ) Outside the state of Pará

##### B. Household insecticide use

4. **Do you use insecticides at home?**

( ) Yes
( ) No

*If NO, end the questionnaire.*

5. **For how long have you been using insecticides?**

( ) < 5 years
( ) 5–10 years
( ) 10–15 years
( ) 15–20 years
( ) 20–25 years
( ) 25–30 years
( ) > 30 years

6. **How often do you use insecticides?**

( ) More than once a week
( ) Once a week
( ) Once a month
( ) Once every 3 months
( ) Once every 6 months
( ) Once a year

7. **Are you the person who applies the insecticide at your home?**

( ) Yes
( ) No

8. **When using insecticides, do you usually wear any protective equipment (mask, gloves, or other)?**

( ) Yes
( ) No
( ) Sometimes

9. **Which type of insecticide do you use most frequently?**

( ) Aerosol spray
( ) Mosquito coil
( ) Bait
( ) Electric device (liquid)
( ) Electric device (tablet)
( ) Other: ________

*Note: in the original field questionnaire, color images illustrating insecticide types were shown to participants to aid recall.*

10. **(Only if "Aerosol spray" was selected in Question 9)** Among the options below, which aerosol brand do you use most often at home?

( ) Brand 1
( ) Brand 2
( ) Brand 3
( ) Brand 4
( ) Other: ________

*Note: in the original questionnaire, color images of the four aerosol brand packages were shown to support correct identification.*
