## Supplementary Table S1 for "Household insecticide use in Amazonian riverine communities: a population-based cross-sectional survey in Belém, Brazil"

**Supplementary Table S2.** Toxicity categories for pyrethroids active ingredients reported in household insecticide products.

| **Active ingredient** | **WHO Hazard Class** | **GHS Category (oral acute toxicity)** |
| --- | --- | --- |
| Cypermethrin | II | Category 3 |
| Deltamethrin | II | Category 3 |
| Esbiothrin | II | Category 4 |
| Imiprothrin | II | Category 4 |
| Prallethrin | II | Category 4 |
| Transfluthrin | U | Category 5 |

Abbreviations: GHS, Globally Harmonized System of Classification and Labelling of Chemicals; U, unlikely to present acute hazard; WHO, World Health Organization.

Active ingredient toxicity was classified according to the WHO Recommended Classification of Pesticides by Hazard and the GHS for oral exposure in acute human health.

**References**

World Health Organization, 2020.WHO recommended classification of pesticides by hazard and guidelines to classification, 2019 edition. World Health Organization, Geneve.
